# Lessons Learned From the Implementation of a Participant-Collected, Mail-Based SARS-CoV-2 Serological Survey in Massachusetts, USA

**DOI:** 10.1101/2021.08.13.21262052

**Authors:** Estee Y. Cramer, Teah Snyder, Johanna Ravenhurst, Andrew A. Lover

## Abstract

The rapid spread of SARS-CoV-2 is largely driven by pre-symptomatic or mildly symptomatic individuals who transmit the virus. Serological tests to identify antibodies against SARS-CoV-2 are an important tool to characterize subclinical infection exposure, which is critical in determining transmission trajectories and consequent population immunity. During the summer of 2020, a mail-based serological survey with self-collected dried blood spot (DBS) samples was implemented among university affiliates and their household members in Massachusetts, USA. Described here are some of the challenges faced and novel procedures used during the implementation of this study to assess the prevalence of SARS-CoV-2 antibodies amid the global pandemic. Important challenges included remote and contact-minimized participant recruitment, limited availability of commodities and laboratory capacity, a potentially biased sample population, and policy changes impacting the distribution of clinical results to study participants. Methods used to surmount these challenges and lessons learned are presented to inform similar studies. Key lessons relate to the acceptability and feasibility of DBS sampling, supply requirements, the logistics of packing and shipping packages, data linkages to enrolled household members, and the utility of having an on-call nurse available for participant concerns during sample collection.

Future studies might consider additional recruitment techniques such as conducting studies during academic semesters when recruiting in a university setting, partnerships with supply and shipping specialists, and using a stratified sampling approach to minimize potential biases in recruitment. This study design highlights the feasibility and acceptability of self-collected bio-samples and has broad applicability for other serological surveys for a range of pathogens.

## Introduction

Serological surveys are an important tool to estimate population-level prevalence of infectious diseases worldwide and are critical for estimating the distribution of SARS-CoV-2 infections.^1–3^ Nearly one-third of people infected with SARS-CoV-2 are asymptomatic^4^; serological surveys can expose underlying transmission dynamics that would otherwise remain hidden due to the prioritization of symptomatic testing in most clinical settings. Therefore, population-based seroprevalence studies directly inform public health decisions by providing data on infection distributions and consequent immunity.

Understanding viral exposure through serological testing is especially useful in university populations where a large percentage of young people may have asymptomatic or subclinical infections. University populations also experience cyclical population mobility due to the academic schedule. Viral transmission among university students may also affect those in close proximity to the university. Though not the case in all large universities and surrounding communities,^5^ it was found that counties with large university populations offering in-person instruction experienced a 56.2% increase in COVID-19 incidence when comparing the 21 days before and after the first day of classes in fall 2020.^6^ These data suggest that viral transmission networks extend beyond universities and into surrounding communities, and may impact vulnerable community members.^7^ This also highlights a need for evidence of transmission for policymakers regarding school reopenings and controlling potential outbreaks.^6,8,9^

Seroprevalence studies benefit from standardized methods, which might include a comparison of the general population with subpopulations, analysis of potential risk factors for infection, and investigation of variations in immune response. A standardized approach is needed to facilitate the comparison of results across studies.^3,10^ Serosurveys of the general population are strengthened with the inclusion of higher-risk subpopulations to help prioritize interventions.^3^ At-home serosurvey sampling using DBS from a mailed testing kit has been validated and found to be a reliable alternative to serum samples for detecting SARS-CoV-2 antibodies.^11,12^ Additionally, there is evidence that collecting venous blood either in-person or at home might be less acceptable to participants than an at-home dried blood spot (DBS) collection.^3,9,10,13,14^ While simpler to use a convenience sample of serology measures, this might reduce validity and generalizability of results. Therefore, many studies employ a population-based random sample and include household members to increase the sample size.^2,3,10^ In addition to testing individuals and their household members for serology results, additional insight regarding demographics and potential risk factors for infection is gained through the combination of serological tests and survey items for data collection.^3,10,13^

During the summer of 2020, there were important considerations to ensure the safety of students returning to campus at the University of Massachusetts Amherst (UMass) due to the COVID-19 pandemic. Research has demonstrated that individuals with asymptomatic or subclinical infections may still transmit SARS-COV-2, potentially contributing to the rapid spread of outbreaks.^4,15^ To better characterize SARS-CoV-2 transmission in MA, a mail-based serosurvey was conducted to quantify the viral exposure among UMass students, faculty, staff, and their household members.

The primary aim of this case study is to describe in detail the implementation of a mail-based serosurvey for the identification of SARS-CoV-2 antibodies in asymptomatic individuals with no prior COVID-19 diagnosis while using participant-collected dried blood spots (DBS). We describe participant recruitment methods, design of the data storage platform, logistics of navigating the shipping supply chain, and statistical analysis. The description of the challenges encountered throughout this study can be used to inform researchers implementing similar mail-based serological studies.

## Methods

### Study Population and Recruitment

The study population included UMass Amherst affiliates (undergraduate students, graduate students, faculty, staff, and librarians) and a single member of their household. UMass affiliates were eligible to participate in the study if they were 18 years of age or older, resided in Massachusetts, reported never having a COVID-19 diagnosis from a healthcare professional, remained in Massachusetts for eight weeks prior to survey administration, and did not report a fever greater than 100.4 □ F at the time of survey completion (Supplemental Table 1). Each UMass affiliate was also asked to invite a member of their household to participate in the study. Eligibility criteria for household members were identical, except household members had to be between 23 and 78 years old to ensure age variation in the population. The restriction to Massachusetts residents who had not left the state was intended to assess the amount of exposure present in The Commonwealth.

UMass affiliates were contacted via email to participate in this study between June 23^rd^, 2020 and June 26^th^, 2020. The invitation email described the study goals, methods, and a unique link to a screening and study eligibility survey. The eligibility survey included an embedded video showing the sample collection process to familiarize participants with blood spot collection. If participants were eligible and agreed to participate, they were asked to electronically sign an informed consent document. Upon consenting, the link opened a detailed survey instrument capturing detailed demographics and potential COVID-19 risk factors (Table 2). The associated household members were also invited to complete these forms themselves.

### Survey Creation and Distribution

All survey instruments were created and administered using Research Electronic Data Capture (REDCap) software version 9.9.0, provided through the University of Massachusetts Medical School. REDCap was chosen for its ability to send unique and HIPAA-compliant survey links to a large number of individuals from an address list. Storing records in this system provided a way to securely link participant demographic information and laboratory results. Email addresses for UMass affiliates were obtained from the University’s Office of Academic Planning and Assessment and were uploaded to REDCap. Emails containing unique survey links for each participant were created in REDCap and sent through the system interface to participants. Emails were sent in batches of several hundred to avoid potential server capacity issues. The survey administered took approximately 5 minutes to complete. Reminder emails were sent to non-respondents 3 and 6 days after the initial email.

After a three-week enrollment period, the survey was locked, and a randomized subset of participants was selected to receive a bio-sample collection kit. Different sampling schemes were used for employees (graduate students, faculty, staff, and librarians) and undergraduate students. The employee sample was generated through a simple random selection of 250 individuals from the eligible population. For undergraduates, 750 individuals were selected based on probability proportional to the population size using the 2019 census data for each of the Massachusetts Emergency Regions.^16^ Emergency regions were used instead of county-level data because sampling at a county level was not feasible. There was not enough representation from all 14 Massachusetts counties, even though many students had returned to their hometowns at the time of survey administration. UMass undergraduate students and their associated family members were referred to as the “primary” sampling group; UMass employees (graduate students, faculty, staff, and librarians) and their family members were the “secondary” sampling group.

### Supply Chains and Shipping Logistics

Each household was sent a single box with supplies for the UMass-affiliated participant and their enrolled household member. While dried blood spots are generally exempt from special mailing considerations, to comply with biosafety considerations during the pandemic boxes appropriate for the transportation of Category B Biological Substances (UN3373)^17^ were used. These were labeled “Biological Substance, Category B, UN3373”, “Exempt Human Specimen.” Bubble wrap was included in the shipping boxes to protect the contents. Box contents contained a bloodspot card with a unique barcode identifier for each participant (stored in bags with silica gel), a sample collection supply bag (gloves, alcohol pads, lancets, bandages, gauze), a biological hazard bag for returning all collection materials, and a detailed pamphlet with sample collection instructions. This pamphlet contained detailed “how-to” information to properly collect the dried bloodspot sample and instructions for properly sealing the biological hazard return bag (Figure 1, Supplemental File 1). After the sample was collected and dried, participants were instructed to place the bloodspot card into the bag with silica. This bag was then placed into the biological hazard return bag, and all box contents were placed into the shipping box for return.

**Figure 1:**
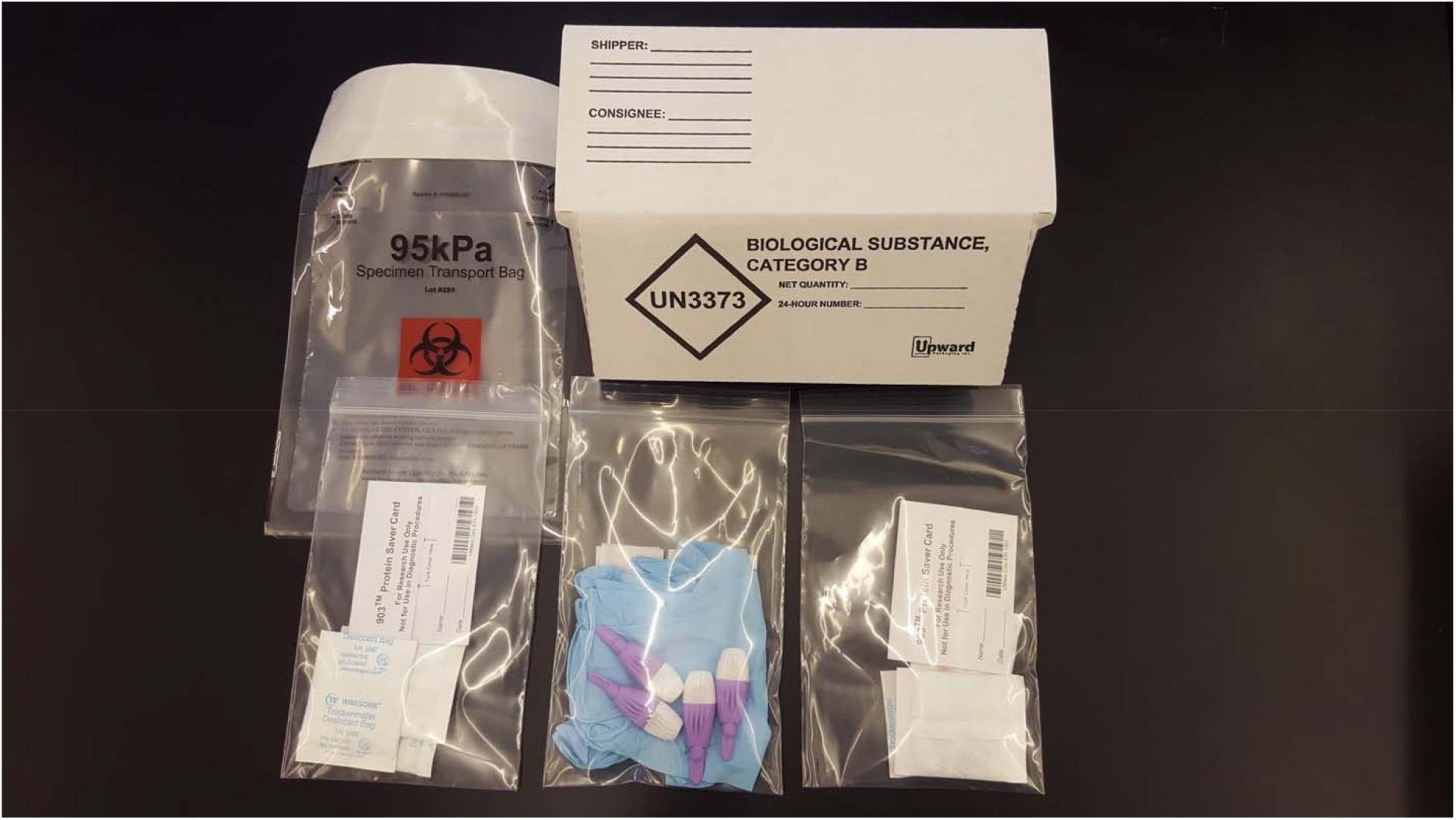
Contents of Shipped Boxes. Each box mailed to a household contained: DBS cards with barcodes, two silica packets per DBS card, collection supplies (gloves, lancets, gauze, alcohol pads, and adhesive bandages), biospecimen bags, return labels, tape to seal the box for return, and instruction sheets for how to collect the sample and how to close the biospecimen bag.

Boxes were shipped to participants using United Parcel Service (UPS) as it was the most economical and simplest for bulk mailings and allowed for tracking to ensure arrival at each participant’s home. United States Postal Service (USPS) was used to deliver to PO boxes, as UPS was unable to deliver to these locations. Generic return labels and shipping tape were included in all kits so the same boxes could be returned via USPS. USPS was used for all returns to make the shipment as simple as possible for participants. Boxes could be returned at any post office, into blue mailboxes, or directly from home mailboxes. UPS was a less feasible option for return shipments, as participants would have needed to locate a UPS location for drop-off, many of which were closed due to the pandemic itself.

### Sample Processing

Returned test kits were processed in a biosafety level 2 laboratory. External box surfaces were sanitized with 70% ethanol before being opened in a HEPA filtered Class II Biosafety Cabinet. All excess supplies were either autoclaved or disposed of in sharps containers as per standard laboratory practice. The bloodspot cards were removed from return bags and matched to individuals in REDCap using the name provided on the outgoing shipping label. Barcodes attached to the DBS cards were then verified with the participant-entered barcodes in REDCap and entered into the second online survey form. Outgoing shipping labels were consulted to confirm and triangulate the barcodes and to link the few participants who did not complete the final online survey signifying that they had received the boxes.

DBS cards were heat-treated at 56 □ C for 30 minutes before bloodspots were manually punched using a 0.25-inch diameter stationery hole puncher. A single bloodspot per participant was transferred to an ELISA plate and assayed for IgM and IgG SARS-CoV-2 antibodies as described in published protocols.^18^ Each 96-well ELISA plate contained one positive control and seven negative controls. Optical densities were read at 405 nm and normalized to the negative controls’ daily average optical density.

### Statistical Analysis

Seropositivity cutoff values were determined using finite mixture models.^19,20^ These models identified the optimal break-point between seronegative and seropositive subpopulations. For this survey, samples with an optical density ≥ 2.49 above daily control values were considered positive for SARS-CoV-2 IgG antibodies. Logistic regression models were used to determine propensity scores for each individual to calculate non-response weights that were applied via inverse weighting. Undergraduate students were self-weighted due to the probability proportional to population size sampling scheme; employees did not require weighting as simple random sampling was used. Poisson models with robust errors to account for household clustering were utilized for separate multivariable regressions within each sampling scheme. Final models were identified using AIC/BIC and were adjusted for age, race, and gender. Analyses were performed with R version 4.0.3^21^ and SAS version 9.4 (SAS Institute, Cary, NC, USA).

## Outcomes

Initial emails were sent to 27,339 UMass affiliates; of the 4,531 individuals who completed the required documents, 407 were ineligible. Of the 4,124 eligible individuals, 1,001 were randomized to receive a test kit (752 undergraduate students and 249 employees). Among the undergraduate students, 330 enrolled a household member; 548 students and 231 household members returned samples and were included in the analysis. Among employees, 101 enrolled a household member; 214 employees and 78 household members returned samples and were included in the analysis. Overall, 76% of randomized participants returned blood samples for analysis (Figure 2).

**Figure 2:**
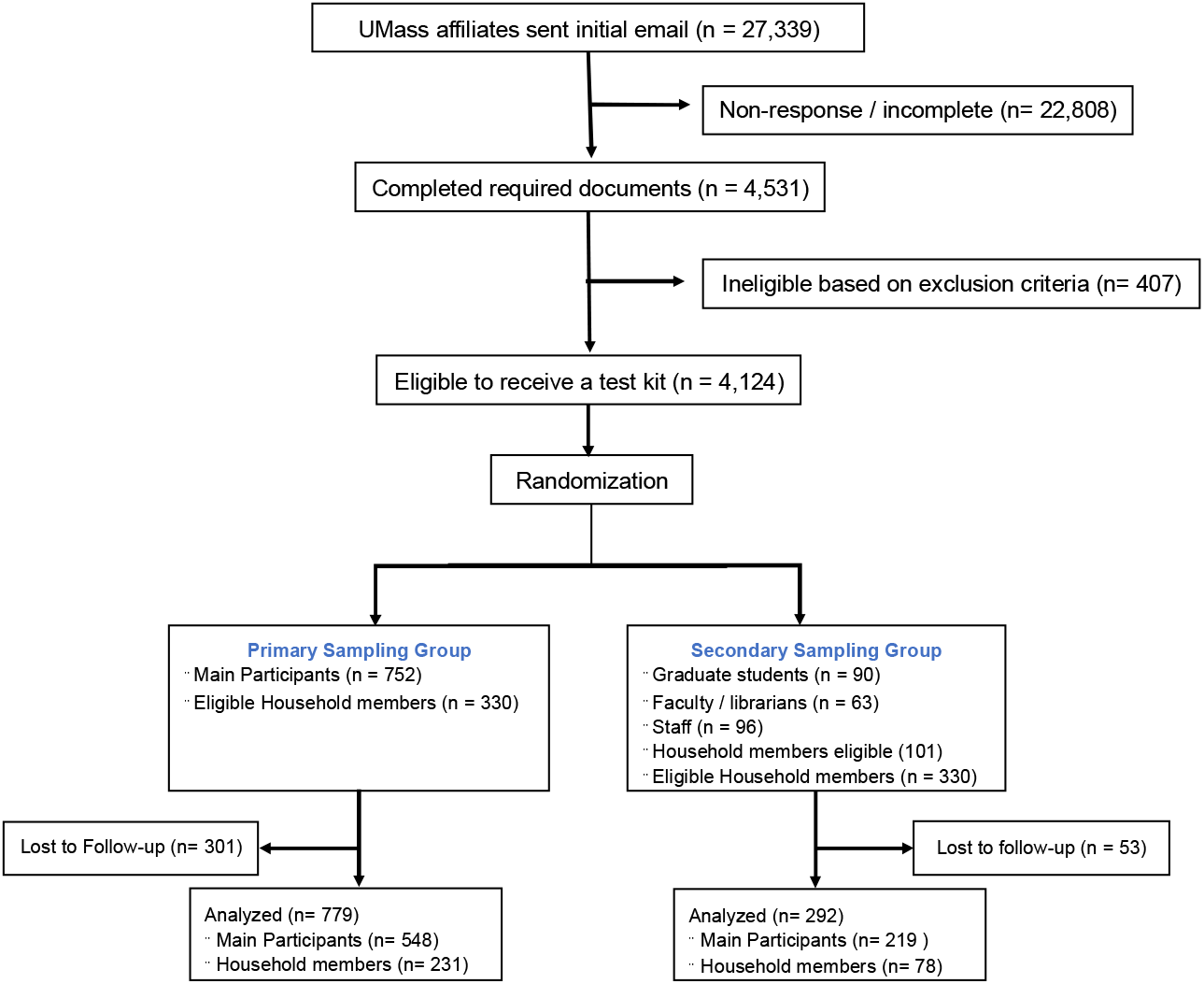
Participant enrollment diagram, SARS-CoV-2 serosurvey, Massachusetts, USA, Jul-Aug 2020

Forty-six samples out of 1,071 total samples were positive for IgG SARS-CoV-2 antibodies. None of the samples tested positive for IgM antibodies. After adjustment for non-response, the overall seroprevalences for participants and their household members was 5.3% (95% CI: 3.1 to 7.5%) for undergraduate students and 4.0% (95% CI: 1.6 to 6.5%) for employees. Factors shown to be significantly associated with seropositivity in the undergraduate analysis included Black or American Indian/Alaska Native race, male gender, and previous febrile illness since February 2020. No geographic locations were associated with seropositivity. Geographic location was significantly associated with seropositivity in the employee analysis, but gender, race, and prior febrile illness were not associated with seropositivity in fully adjusted models.^22^

## Lessons Learned

### Supply Acquisition and Staffing Considerations

The acquisition of supplies was unexpectedly complex due to nationwide supply shortages, shipping delays, and university staff furloughs due to the pandemic. Routine items, including alcohol pads, sterile gauze, medical gloves, and biological sample shipping boxes were back-ordered or completely unavailable with some suppliers. Using a Canadian supplier for shipping boxes added additional administrative processes (which were complicated by university furloughs) and resulted in delays due to customs clearances. Supply shortages for some supplies (alcohol pads and sterile gauze) were remedied with large purchases at local pharmacies, while others (medical gloves) were obtained from university affiliates. Moreover, research restrictions enacted by the university severely limited room occupancies throughout campus. To comply with these spacings, two graduate students received clearance from the university and created an assembly line to pack boxes and test samples; larger staffing would have greatly decreased the overall time required for this process.

After sending out the first batch of approximately 100 boxes, participants reported that the lancet blade was too small to obtain sufficient blood flow, making it difficult or impossible to collect bloodspots. Additionally, as all lancets self-retracted after a single use, some participants used all that were sent as they were unfamiliar with the design. This situation required repackaging the remaining boxes with larger bladed lancets and shipping new lancets to the first batch of participants. Ensuring that the types of lancets chosen have large enough blades to obtain the desired sample is essential (e.g., 1.5mm bladed, with 1.6mm penetration depth), and several extras should be included. Future studies should consider pilot shipments of a few kits to participants to collect feedback before sending out the remaining test kits, which may prevent the need to ship out new supplies to all participants if they are determined to be inadequate.

### Coordination with Mail Services

Close coordination with the UMass Campus Mail Services was essential in dealing with the challenges inherent in shipping approximately 1,000 boxes. Outgoing boxes were delivered to the University loading dock over several days; special pickups were coordinated with UPS due to the size of the shipments. Mail Services added extra staff and additional deliveries to accommodate the large volume of returned boxes. Due to shipping delays and backlogs with UPS and USPS, several boxes were delivered to participants much later than anticipated. Participants were granted extended deadlines and could return boxes to the laboratory in person to maximize sample collection. Future studies that use at-home test kits should prioritize collaboration with shipping specialists and should anticipate possible delays at all stages of the shipping process.

### Response Rates

When conducting this survey, there were concerns that many university students might not check their institutional emails over the summer months. Overall, the non-response rate for the study was 83.4% among UMass affiliates. Although the survey completion rate appears low, it is slightly higher than other studies related to COVID-19 conducted on large university college campuses during 2020.^23–25^ Therefore, it seems that UMass affiliates continued to monitor their academic emails over the summer vacation period.

In this survey, attempts to improve participation rates were made by keeping the surveys short (∼5 minutes), keeping the enrollment period open for three weeks, and sending reminder emails to non-respondents 3 days and 6 days after the initial email. Additionally, attempts to increase participation included informing participants about the length of the survey and giving them the option to receive their serological results upon completion of the study. To achieve higher response rates, future studies should consider sending out the survey from a more familiar office on campus, such as the office of the dean.^26^ This could help ensure that emails receive proper attention and aren’t sent to “Junk” folders.

One reason for concern regarding low response rates is the potential of selection bias due to the types of individuals most likely to respond to volunteer-based surveys.^27^ Although it is difficult to quantify the extent to which selection bias may have impacted findings, among the primary sampling group, a larger percentage of women responded (62%) compared to the overall proportion of undergraduate women at UMass (51%). There was also slightly less representation of minority populations (African American, Chicano/Latino, and Native American/Alaska Native) in the study population than in the broader UMass undergraduate population. In parallel, among the secondary sampling group, there was a higher percentage of White and Asian individuals, but there was not a difference in the percentage of females relative to the overall demographics.^28^ Men may be more susceptible to SARS-CoV-2 infection and fatality^29,30^ so this overrepresentation of women may have underestimated the SARS-CoV-2 exposures present in the population. Similarly, during the summer of 2020, minority groups were more likely to contract SARS-CoV-2,^31^ therefore, the overrepresentation of White and Asian race categories may have underestimated the overall exposures in this sub-population. In survey studies, it is common that women and individuals with lower risk factors are more likely to participate,^27^ so future studies should consider oversampling men and minority groups to better capture population-level prevalence.

### Randomization

After the three-week enrollment period, randomization of participants to receive an at-home test kit occurred for all eligible participants to avoid choosing the first group of individuals to enroll in the study. These individuals could have been more motivated to participate due to known exposure or other factors. In this study, two sampling approaches were used. For the primary study population, a stratified sampling approach was used to capture a sample representative of individuals from across the state of MA. For the secondary sample, a simple random sample was used. Future studies may consider a more stratified sampling approach to better capture the diversity in race and gender and thus collect a more representative sample of the broader population.

### Survey Questions

A probable source of drop-out from household members who initiated the survey but did not complete it was confusion over a survey eligibility question regarding UMass affiliation. In the eligibility screen, the question was, “Is someone **in your household** a current UMass student or employee?” Some people were confused and thus answered incorrectly and were deemed ineligible to complete the survey and be included in testing. Given the timing of these surveys, it was not possible to do a large-scale pilot test. Future studies should include as large a pilot as possible.

Additionally, after sending out test kits, individuals were sent a brief form asking them to enter their barcode information into the web portal from the barcoded blood spot card (as it was not feasible to allocate the > 1000 boxed codes to individual addresses). Unfortunately, a number of participants returned their kits without completing this form, thus making it difficult to match samples to individuals. A potential solution would be to record the barcode of each test strip mailed to each individual before mailing, or to include a physical note in the test kit box reminding all recipients to complete the online form.

### Result Distribution

A major unanticipated obstacle occurred when the guidance surrounding the distribution of COVID-19 related results to participants was clarified. With approval from the IRB, participants were offered the option to receive their individual test results when initially signing up for the study. A few months after this determination, CDC guidance was expanded to cover reporting of all SARS-CoV-2 test results, which meant that any analyses performed in a laboratory setting that was not CLIA-certified could not be disseminated to participants. Due to these changes, only aggregate results could be shared with study participants instead of individual results. Samples are being retested in a CLIA-certified laboratory at UMass so that participants can obtain their results.

## Data Availability

Data are available upon reasonable request.

## Summary

Despite the challenges encountered throughout implementing this study, the methods used for sample collection were shown to be efficient and effective for obtaining contact-free seroprevalence data by mail. This study used novel strategies for its study design by selecting a subset of university affiliates, enrolling their household members, and utilizing participant-collected blood samples using at-home DBS kits. Administering online questionnaires to UMass affiliates and their family members was a rapid method to obtain a representative sample of Massachusetts residents’ potential SARS-CoV-2 antibody statuses. This mail-based, minimal contact study was a safe, cost-effective, and pragmatic way to assess prior infection with SARS-CoV-2 during the height of the pandemic.

When conducting future studies relying on the at-home collection of samples, consideration should be given to the availability of shipping supplies, potential complexities with shipping processes, methods to increase response rates among eligible participants, and determining how regulatory changes may impact roll-out and reporting (Table 1). This report highlights the feasibility and acceptability of self-collected biosamples and has broad applicability for other serological surveys for a wide range of other infectious diseases.

**Table 1.**
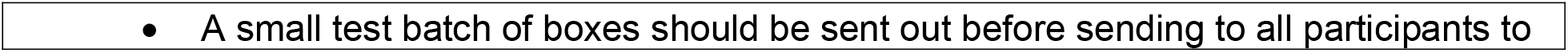
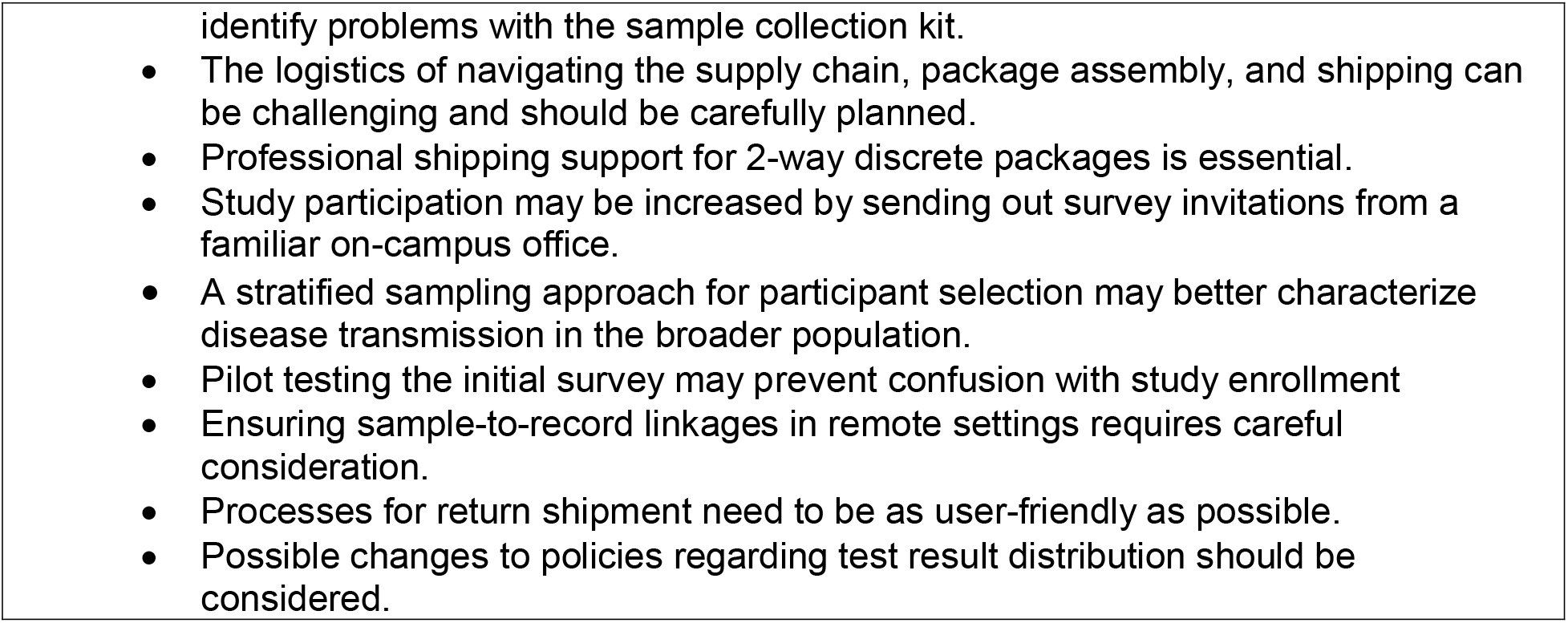
Lessons learned from the implementation of a mail-based SARS-CoV-2 serosurvey.

## Supplemental Files

**Supplemental Table 1:**
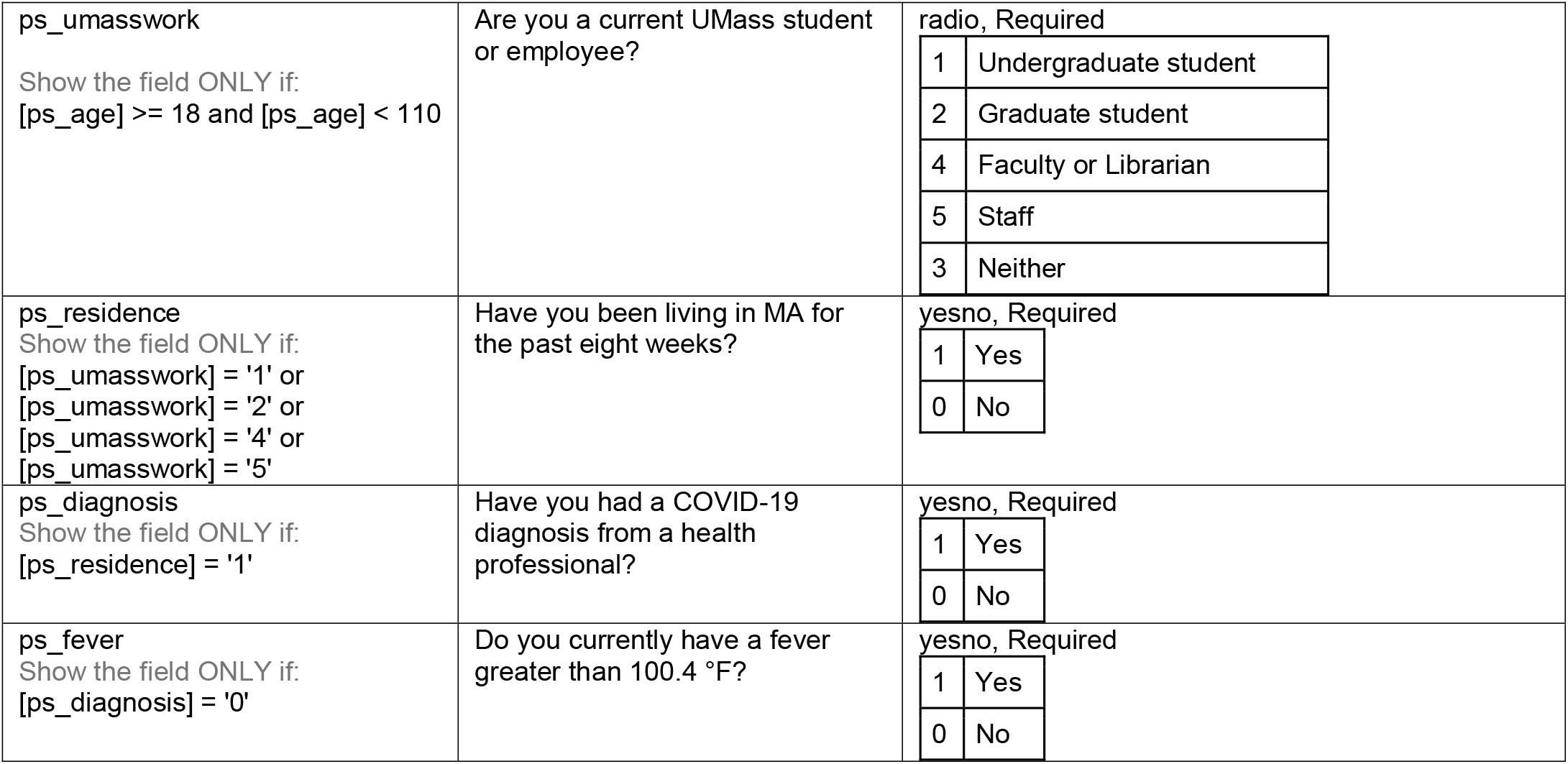
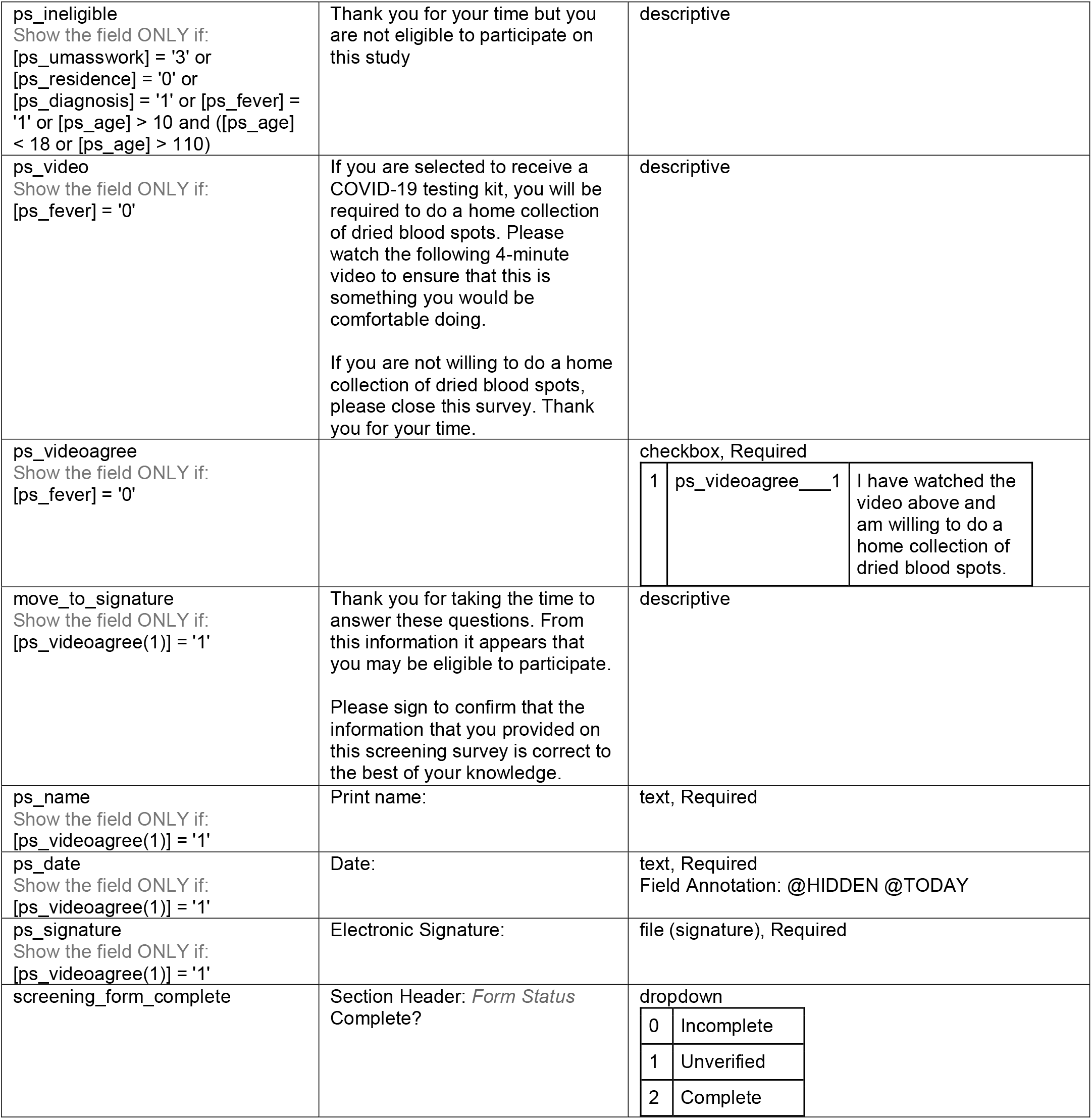
Prescreen survey codebook

**Supplemental Table 2:**
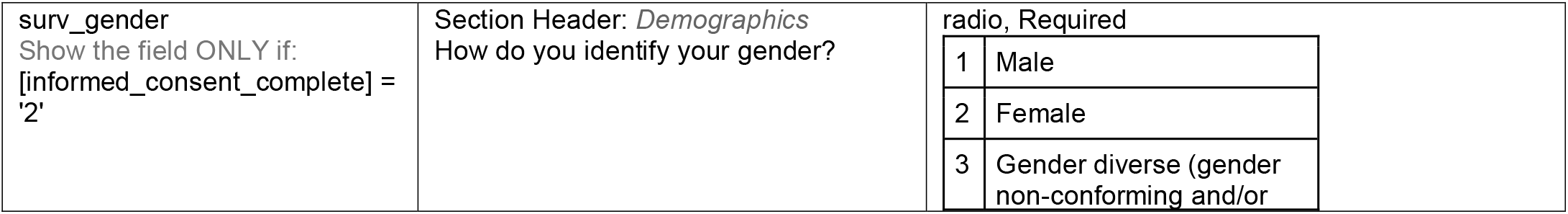
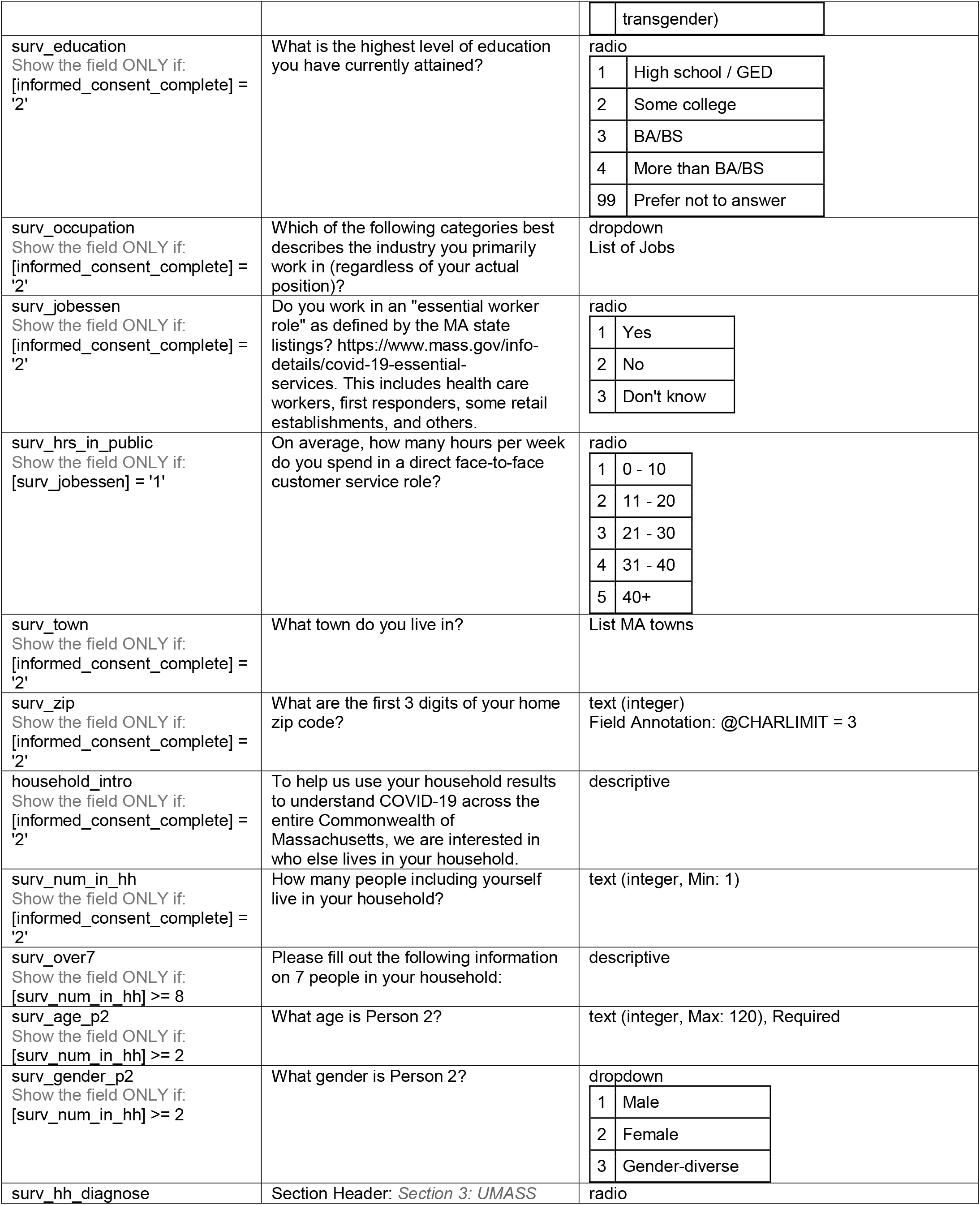
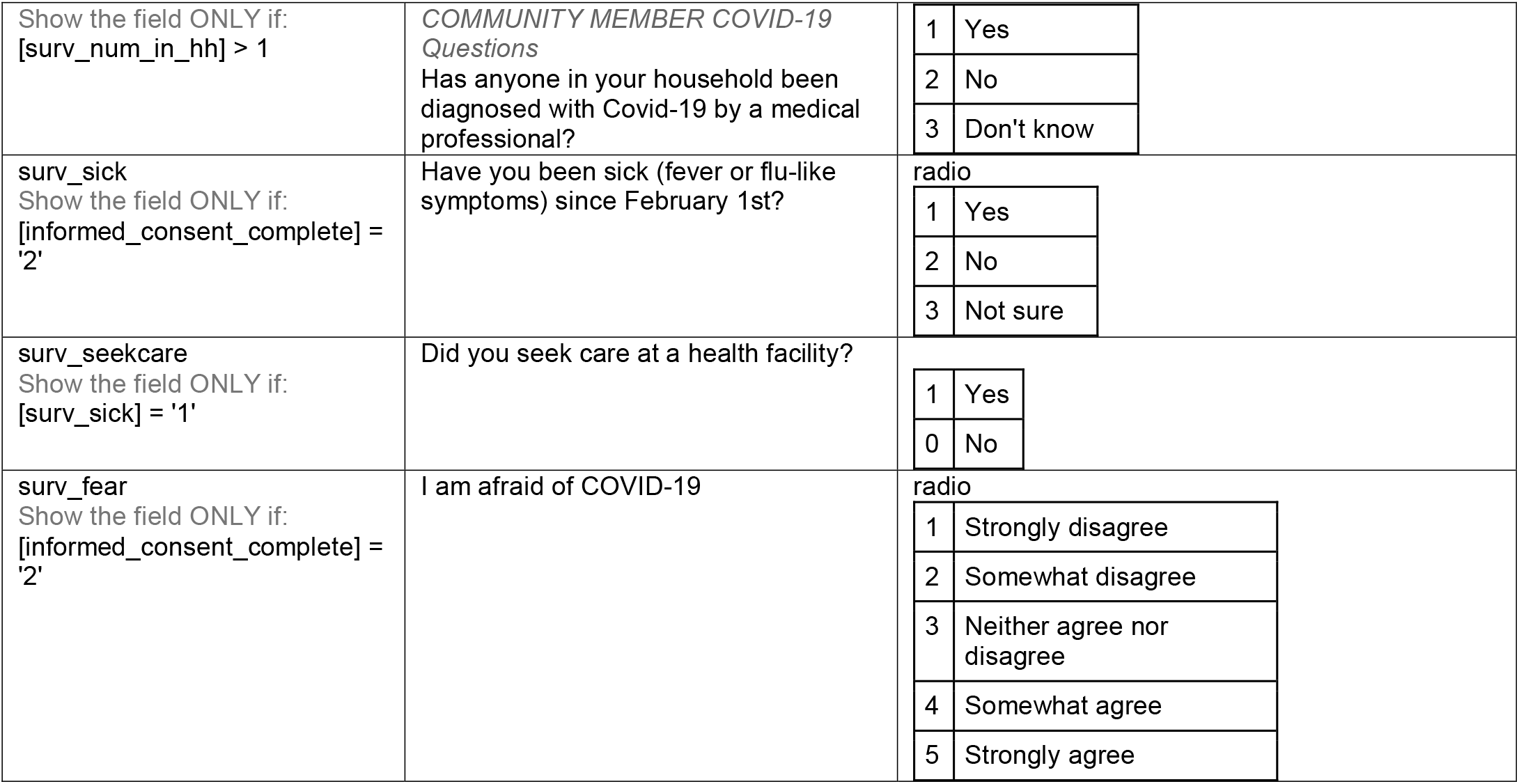
Survey questions asked to each UMass affiliate

**Supplemental File 1:**
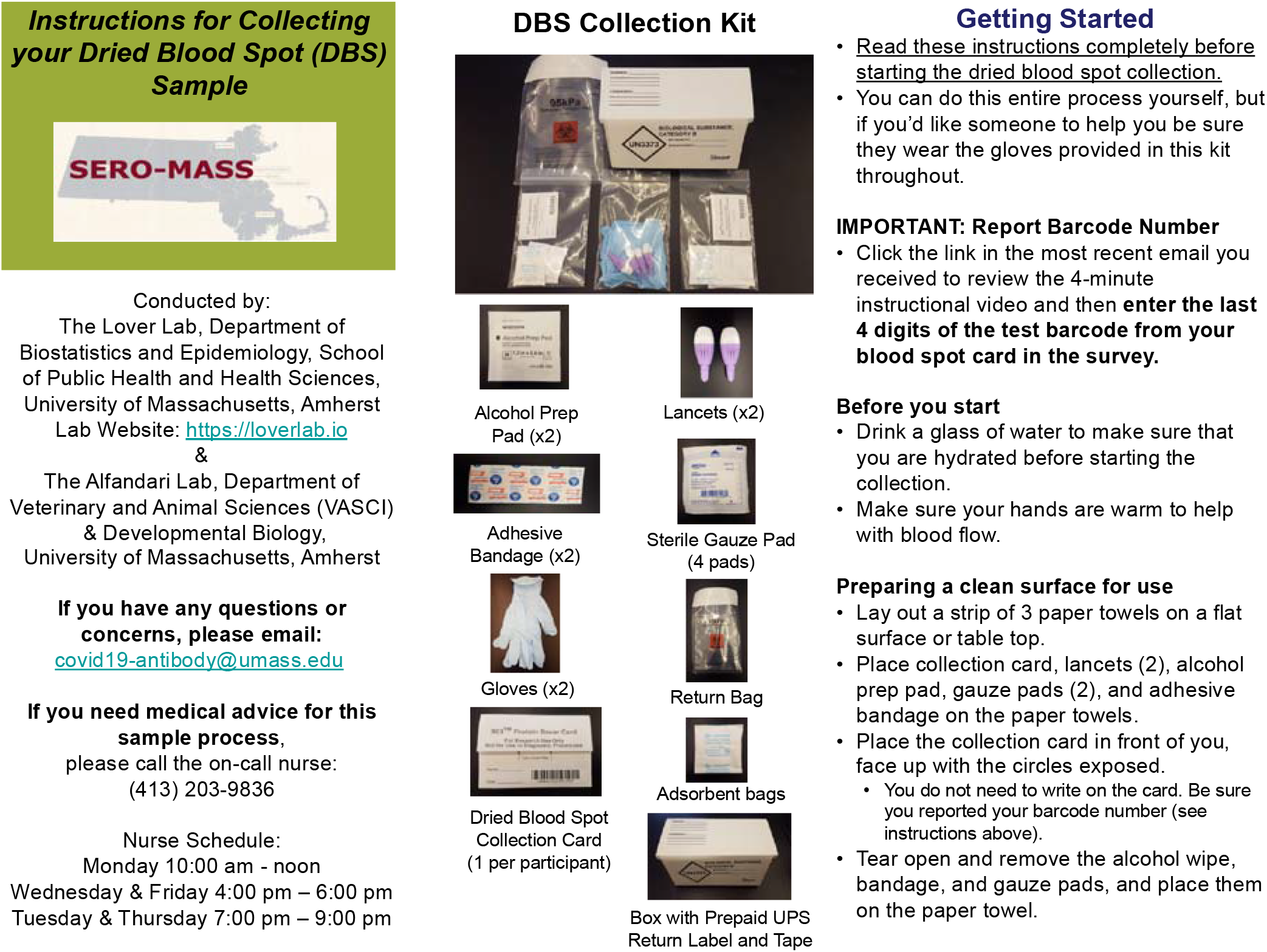
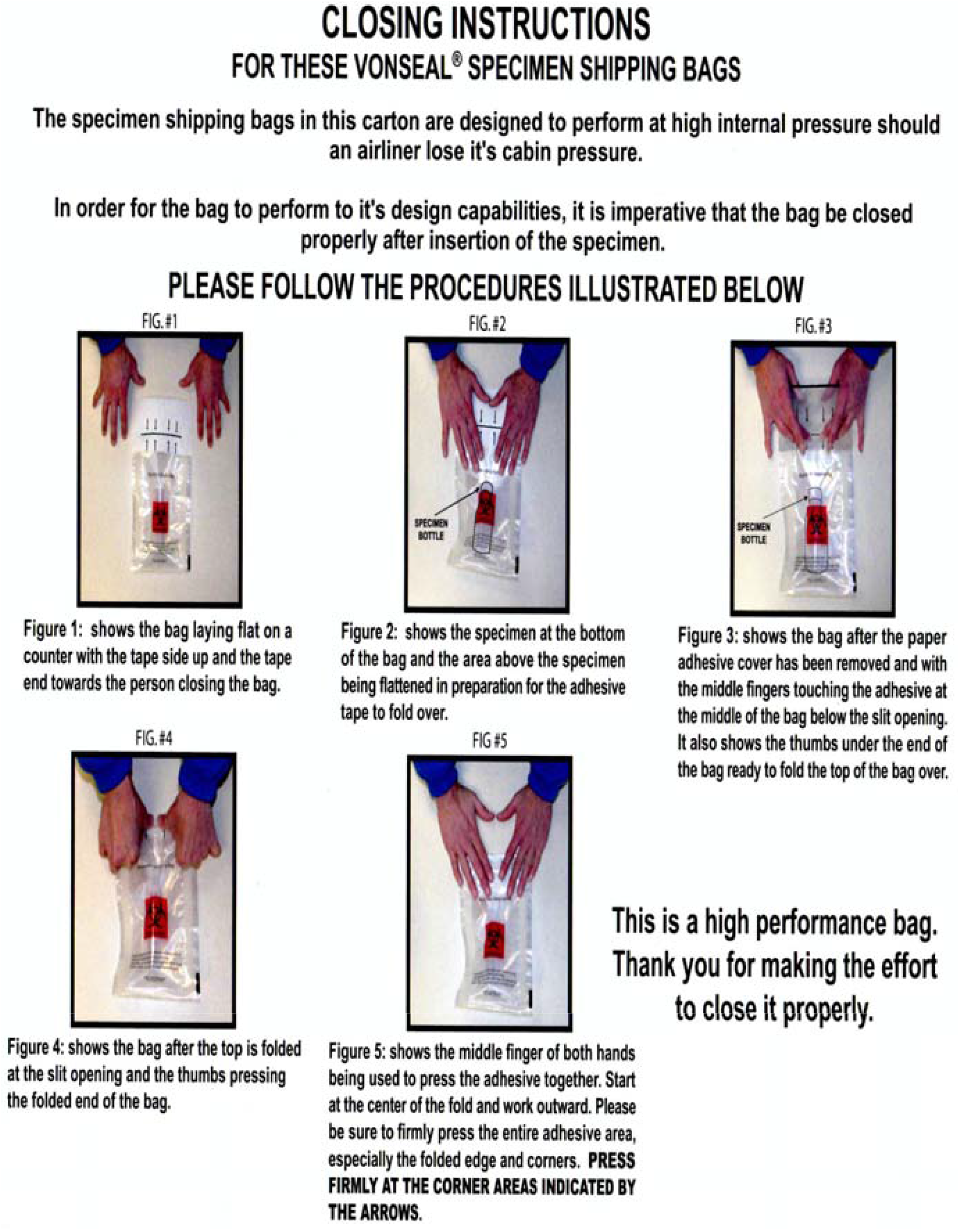
Instructional guides for collecting and repackaging samples. PDF versions of files were included with every box giving users instructions on collecting blood spots and properly mailing back the packages.

## Notes

### Competing Interest Statement

The authors have declared no competing interest.

### Funding Statement

UMass Faculty Funds (A Lover; SPH-AL-001); UMass Faculty Funds
(N Reich; SPH-NR-001); UMass Institute for Applied Life Science 'Midigrant' (#169076; A Lover) and D Alfandari was supported by grants from the NIH/USPHS (RO1DE016289 and R24OD021485).

### Author Declarations

This study was approved by the University of Massachusetts- Amherst Human Research Protection Office (Approval #2062; 27 Apr 2020).

